# Sensitivity of rapid antigen testing and RT-PCR performed on nasopharyngeal swabs versus saliva samples in COVID-19 hospitalized patients: results of a prospective comparative trial (RESTART)

**DOI:** 10.1101/2021.04.09.21255105

**Authors:** Antonios Kritikos, Giorgia Caruana, René Brouillet, John-Paul Miroz, Abed-Maillard Samia, Stieger Geraldine, Onya Opota, Antony Croxatto, Peter Vollenweider, Pierre-Alexandre Bart, Jean-Daniel Chiche, Gilbert Greub

**Author notes:** **Corresponding Author:** Prof. Gilbert Greub, Institute of Microbiology, Department of Laboratory Medicine, Lausanne University Hospital and University of Lausanne, Rue du Bugnon 48, CH-1011, Lausanne, Switzerland. Personal. Equal contribution (Antonios Kritikos, Giorgia Caruana).

## Abstract

**Objectives:** Saliva sampling could serve as an alternative non-invasive sample for SARS-CoV-2 diagnosis while rapid antigen testing (RAT) might help to mitigate the shortage of reagents sporadically encountered with RT-PCR. Thus, in the RESTART study we compared antigen and RT-PCR testing methods on nasopharyngeal (NP) swabs and salivary samples.

**Methods:** We conducted a prospective observational study among COVID-19 hospitalized patients between 10^th^ December 2020 and 1^st^ February 2021. Paired saliva and NP samples were investigated by RT-PCR (Cobas 6800, Roche-Switzerland) and by two rapid antigen tests: One Step Immunoassay Exdia^®^ COVID-19 Ag (Precision Biosensor, Korea) and Standard Q^®^ COVID-19 Rapid Antigen Test (Roche-Switzerland).

**Results:** A total of 58 paired NP-saliva specimens were collected. Thirty-two of 58 (55%) patients were hospitalized in the intensive care unit and the median duration of symptoms was 11 days (IQR 5-19). NP and salivary RT-PCR exhibited sensitivity of 98% and 69% respectively whereas the specificity of these RT-PCRs assays were of 100%. NP RAT exhibited much lower diagnostic performances with sensitivities of 35% and 41% for the Standard Q^®^ and Exdia^®^ assays respectively, when a wet-swab approach was used (i.e. when the swab was diluted in the viral transport medium (VTM) before testing). The sensitivity of the dry-swab approach was slightly better (47%). These antigen tests exhibited very low sensitivity (4 and 8%) when applied to salivary swabs.

**Conclusions:** Nasopharyngeal RT-PCR is the most accurate test for COVID-19 diagnosis in hospitalized patients. RT-PCR on salivary samples may be used when nasopharyngeal swabs are contraindicated. RAT are not appropriate for hospitalized patients.

## Introduction

Rapid and accurate detection of SARS-CoV-2 infection in hospitalized patients is the cornerstone of prompt patient care and contact tracing. To date, nasopharyngeal (NP) swab real-time polymerase chain reaction (RT-PCR) remains the reference specimen for SARS-CoV-2 testing (1). However, NP swabing might expose healthcare workers to the risk of transmission during sampling and is a relatively invasive method, especially when considering the multiple samplings a patient will go through during his hospital stay. On the other hand, there is growing evidence advocating on the role of salivary or oropharyngeal specimens as alternative non-invasive methods for SARS-CoV-2 diagnosis (2-6).

At the same time and as a response to the growing SARS-CoV-2 pandemic and reagents shortages for rapid molecular systems, multiple rapid point-of-care tests (POCTs) have been added to the diagnostic pipeline for COVID-19 (7, 8).

Little is known on the utility and diagnostic performances of the above-mentioned strategies in moderately to severely ill hospitalized patients. Current literature shows discordant results, depending on the clinical setting studied, the sampling method or even the use or not of a viral transport medium (VTM) (2-6, 9-17).

In order to simultaneously investigate analytical [RT-PCR versus Rapid Antigen Test (RAT)] and sampling procedures (NP swab versus saliva specimen and use versus not of VTM) we conducted a prospective observational study in hospitalized patients. Our aim was to:

1. compare the diagnostic performances of NP RT-PCR and salivary RT-PCR,
2. evaluate the reliability of RAT in hospitalized patients,
3. compare the sensitivity of RATs performed on NP swab versus saliva in this specific population,
4. evaluate the impact of VTM on the diagnostic performance of RAT.

## Methods

### Study population

We conducted a prospective observational study among SARS-CoV-2 positive patients hospitalized in our institution, a tertiary university hospital in Lausanne, Switzerland (CHUV), between December 10, 2020 and February 1, 2021. All patients were previously COVID-19 confirmed cases (via NP RT-PCR) and hospitalized either in internal medicine ward, or in intensive care unit (ICU). Inclusion criteria were a) positive NP RT-PCR for SARS-CoV-2 in the previous 5 days, b) age >18 years-old and c) informed consent acquisition by the patient or the next of a kin for patients incapable to provide informed consent. We collected no additional personal or clinical data beyond the usual information required for every SARS-CoV-2 test by the Federal Office of Public Health (FOPH) and our microbiology laboratory (age, sex, hospitalization ward, type and duration of symptoms).

### Sample Collection and Diagnostic tests

After informed consent acquisition, patients underwent two NP swabs [one diluted in universal VTM (UTM^®^ Copan Diagnostics) (“Wet” approach) (17) and one non-diluted (“Dry approach”)] and a saliva sample also diluted in the same VTM. Paired diluted NP and saliva samples were analyzed by RT-PCR (Cobas 6800^®^, Roche-Switzerland) (18) and by two rapid antigen tests (RAT): One Step Immunoassay Exdia COVID-19 Ag (Precision Biosensor, Korea) and Standard Q^®^ COVID-19 Rapid Antigen Test (Roche-Switzerland). The non-diluted NP swab was tested with Standard Q^®^ COVID-19 Rapid Antigen Test (Roche – Switzerland) at patient’s bedside in order to evaluate the effect of VTM on diagnostic performances of RAT. Both “Wet” and “Dry” approaches are recommended by the manufacturers of both RAT used. An additional manipulation step using VTM tubes was performed for an in vitro evaluation of a possible dilution effect of VTM (see section ***“****In vitro testing of dilution effect” below*).

All samples were taken by two specialists in infectious diseases (AK and GC) or a member of a paramedic team trained in NP swabs collection (coming from a team performing all COVID-19 samples in our hospital). NP was performed according to the recent CDC and WHO guidelines (19, 20) and saliva sampling protocol was based on previously published data adapted for hospitalized patients (**supplementary material, S1**) (5). Each nasopharyngeal swab was performed on a different naris, which was randomly chosen for each sample.

### Quantitative SARS-CoV-2 PCR and RAT

RT-PCR was performed in our microbiology laboratory using the automated Cobas 6800^®^ system (Roche-Switzerland) (18). In order to quantify the viral load (VL) based on the number of cycles threshold (Ct) obtained with the molecular platform, we used the following equation, derived from RNA quantification: VL = (10^((Ct −40.856)/ −3.697))*100. Details on methods used to derive this equation were described elsewhere (21). The analytical limit of detection was determined to be at 1000 copies per ml. For graphical representation purposes, NP or salivary samples with undetectable VL are represented in graphs with VL determined to be at 500 copies/ml.

NP swabs and saliva samples were used to assess RAT performances. NP swabs were either directly suspended in the buffer solution (“Dry approach”), or initially suspended in 3 ml of VTM (“Wet” approach). One hundred and fifty µl of the sample were subsequently mixed with the buffer solution. Saliva samples were only treated with the “Wet” approach. Reading of the results was performed after 15 to 30 minutes as specified by manufacturer’s instructions (on a band of immunochromatography paper for Standard Q^®^ COVID-19 Rapid Antigen Test or using Exdia TRF Biosensor to detect immunofluorescence signal for One Step Immunoassay Exdia COVID-19 Ag). In case of doubtful results, the Standard Q^®^ COVID-19 Rapid Antigen Test test was read independently by a second person.

### In vitro testing of dilution effect

In order to test *in vitro* a possible dilution effect generated by the use of VTM tubes instead of direct testing, we simulated the two sampling scenarios: the “Wet” scenario versus the “Dry” swab scenario (**supplementary material, S2_Figure 1**). A SARS-CoV-2 positive clinical NP sample was diluted 7 times and the series was used as internal reference for the limit of detection. Each initial sample of this dilution series represented a “Dry” swab. Then, two clean swabs were soaked in each one of the dry samples and then suspended in two further VTM tubes, thus simulating two series of “Wet” samples. Each sample from every series was tested as follows:

i. One Step Immunoassay Exdia COVID-19 Ag (Precision Biosensor, Korea) and Standard Q^®^ COVID-19 Rapid Antigen Test (Roche-Switzerland) were performed after mixing 350 µl of VTM with the buffer solution provided in the RAT kit;
ii. a clean swab was first inoculated in the VTM, then mixed with the buffer solution (according to manufacturer instructions) and tested with the previously mentioned commercial kits and
iii. 300 µl of VTM were used for RT-PCR analysis.

### Statistical analysis

Descriptive statistics were presented as number and percentage for categorical variables and mean ± standard deviation (SD) or median (interquartile range; IQR) for continuous variables. Chi-square or Fisher’s exact test were used for categorical variables and Wilcoxon matched-pairs rank test for continuous variables where appropriate. Sensitivity, specificity, positive and negative predictive values and 95% CI were calculated to assess diagnostic performances using a positive NP or saliva RT-PCR as reference standard. All statistical analyses were performed using GraphPad Prism version 8.3.0 for Windows (GraphPad Software, San Diego, California USA).

### Ethics

This project was conducted in accordance with the Declaration of Helsinki, the principles of Good Clinical Practice and the Swiss Human Research Act (HRO). The project received approval from the Ethics Committee of canton Vaud, Switzerland (2020–02818). The study was registered on ClinicalTrials.gov with the number NCT04839094.

## Results

### Patients’ characteristics

All patients with confirmed SARS-CoV-2 infection by NP RT-PCR and admitted in ICU or Internal Medicine ward during the study period were screened for eligibility criteria. **Figure 1** shows the flowchart of the screening process. A total of 58 paired NP and saliva samples were performed with a positivity rate of 85% (time elapsed from screening sample to inclusion could be up to 5 days). Baseline demographics and clinical characteristics are shown on **Table 1**. Patients were predominantly males (n=45, 77%) with a median age of 70 years-old (IQR 61-77). Most of them still had symptoms upon sampling (n=49, 84%) with a median duration of symptoms of 11 days (IQR, 5-19). Common symptoms on presentation were dyspnea (n=27, 46%), cough (n=19, 33%) and fever (n=10, 17%). SARS-CoV-2 VL in NP swab ranged from 3.800 to 9.900.000 copies/ml (median value 48.000 copies/ml).

**Table 1:** Baseline demographics and clinical characteristics of patients.

| Characteristics | Internal Medicine Patients<br>(n=26) | ICU patients<br>(n=32) | Overall<br>(n=58) |
| --- | --- | --- | --- |
| Age (y), median (IQR) | 70 (61-77) | 71 (62-78) | 70 (61-77) |
| Male, n (%) | 20 (77) | 25 (78) | 45 (77) |
| Patients with symptoms upon sampling, n (%) | 21 (81) | 28 (87) | 49 (84) |
| Duration of symptoms, median (IQR) | 11 (5-19) | 11 (6-19) | 11 (5-19) |
| Type of symptoms, n (%) |  |  |  |
| Fever | 5 (19) | 5 (16) | 10 (17) |
| Cough | 8 (31) | 11 (34) | 19 (33) |
| Dyspnea | 7 (27) | 20 (62) | 27 (46) |
| Fatigue | 4 (15) | 3 (9) | 7 (12) |
| Anosmia/Dysgeusia | 1 (4) | 2 (6) | 3 (5) |
| Other | 6 (23) | 8 (25) | 14 (24) |
| Viral load, median (IQR) | 43000 (3700-9100000) | 82000 (4500-12000000) | 48000 (3800-9900000) |

(«Viral load» refers to NP swab viral load and is expressed in copies/ml, y=years, IQR=interquartile range).

**Figure 1:**
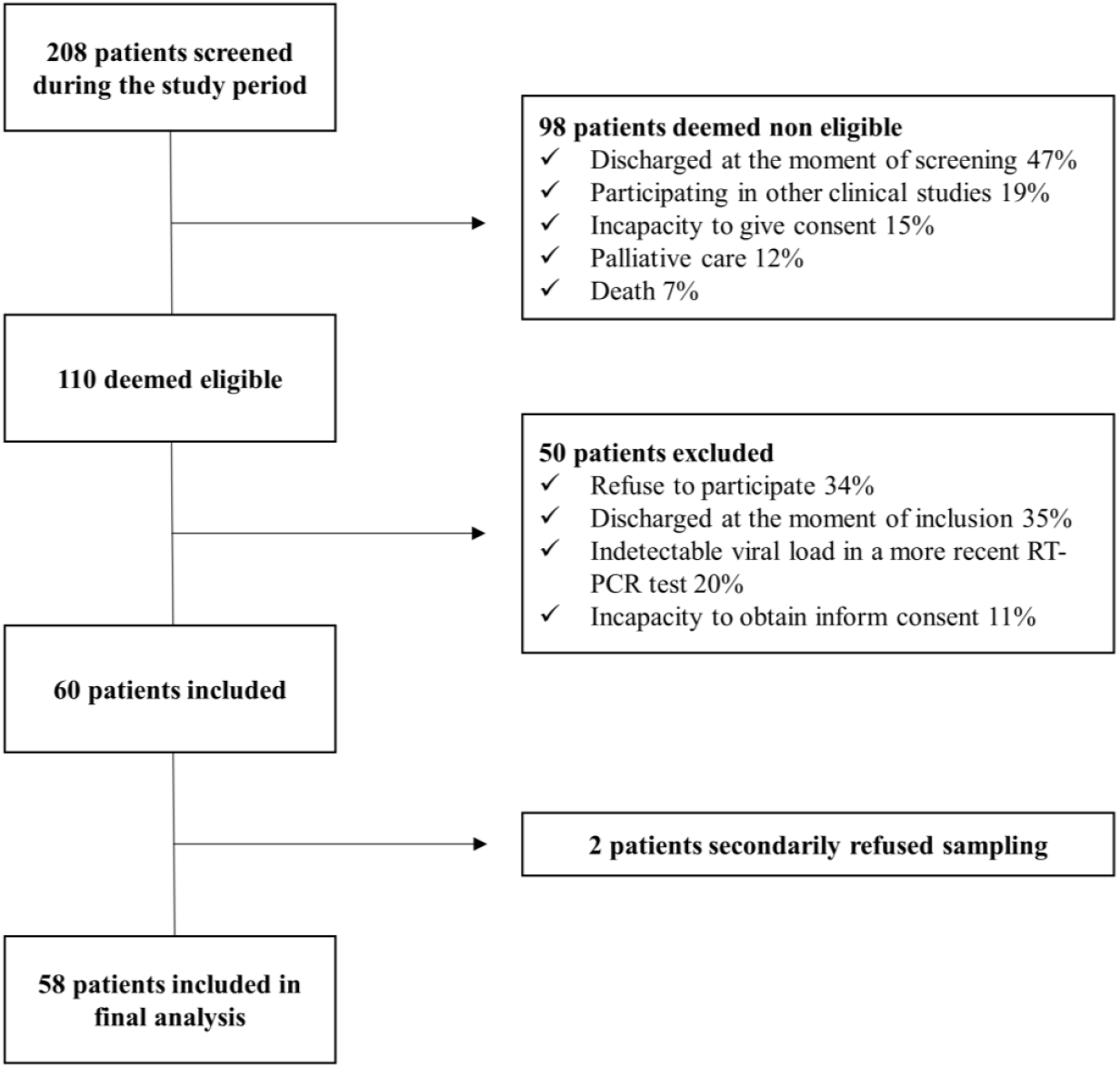
Flowchart of screening process.

### Diagnostic performance of RT-PCR (NP versus saliva) and rapid antigen testing

The diagnostic performance of RT-PCR and RAT for diagnosis of SARS-CoV-2 infection is shown on **Table 2**. A positive NP RT-PCR or salivary RT-PCR was selected as the reference standard comparator.

**Table 2:** Overall diagnostic performance metrics of the different diagnostic approaches used.

| Diagnostic test |  | Sensitivity (%) [95% CI] | Specificity (%) [95% CI] | PPV (%) [95% CI] | NPV (%) [95% CI] |
| --- | --- | --- | --- | --- | --- |
| Nasopharyngeal RT-PCR |  | 98 [89-100] | 100 [66-100] | 100 [93-100] | 90 [56-98] |
| Salivary RT-PCR PCR |  | 69 [55-82] | 100 [66-100] | 100 [90-100] | 37.5 [28-48] |
| Nasopharyngeal rapid antigen test | Exdia®<br>("Wet") | 41 [27-56] | 100 [63-100] | 100 [83-100] | 22 [18-26] |
|  | Roche®<br>("Wet") | 35 [22-50] | 100 [66-100] | 100 [82-100] | 22 [19-26] |
|  | Roche®<br>("Dry") | 47 [35-62] | 100 [66-100] | 100 [86-100] | 26 [21-31] |
| Salivary rapid antigen test | Exdia®<br>("Wet") | 8 [2-20] | 100 [66-100] | 100 [51-100] | 17 [15-18] |
|  | Roche®<br>("Wet") | 4 [0.5-14] | 100 [66-100] | 100 [18-100] | 16 [15-17] |

A positive nasopharyngeal or salivary RT-PCR was used as reference standard. "Wet" and "Dry" rapid antigen tests refer to testing performed after soak of the swab ("Wet") in universal viral transport medium (Copan Italia UTM-RT) or directly to patient's bedside after sampling ("Dry").

NP and salivary RT-PCR exhibited an overall sensitivity of 98% and 69% respectively whereas the specificity of both assays was of 100%. Noteworthy, the sensitivity of salivary PCR increased to 81 % (95% CI: 59-88) in patients presented with a duration of symptoms of less than 10 days. VL (copies/ml) in NP swabs was significantly higher than that detected on salivary specimens for up to 20 days after illness onset (**Figure 2**). Median VL value in positive NP swabs with negative paired saliva specimens was 3700 copies/ml (IQR, 2900-9675). Median duration of illness for those patients was 15 days (IQR, 9-21). ICU patients had higher VL compared to patients hospitalized in internal medicine ward. Pair testing results are shown in **Figure 3**. An analysis of the agreement between the two specimens (NP versus saliva) revealed a fair agreement with a kappa coefficient of 0.37 (95% CI 0.16-0.59; p=0.001) and an overall proportion of agreement of 72% (proportion of positive agreement 80% and proportion of negative agreement 53%) (**supplementary material, S3_Figure 2**).

**Figure 2:**
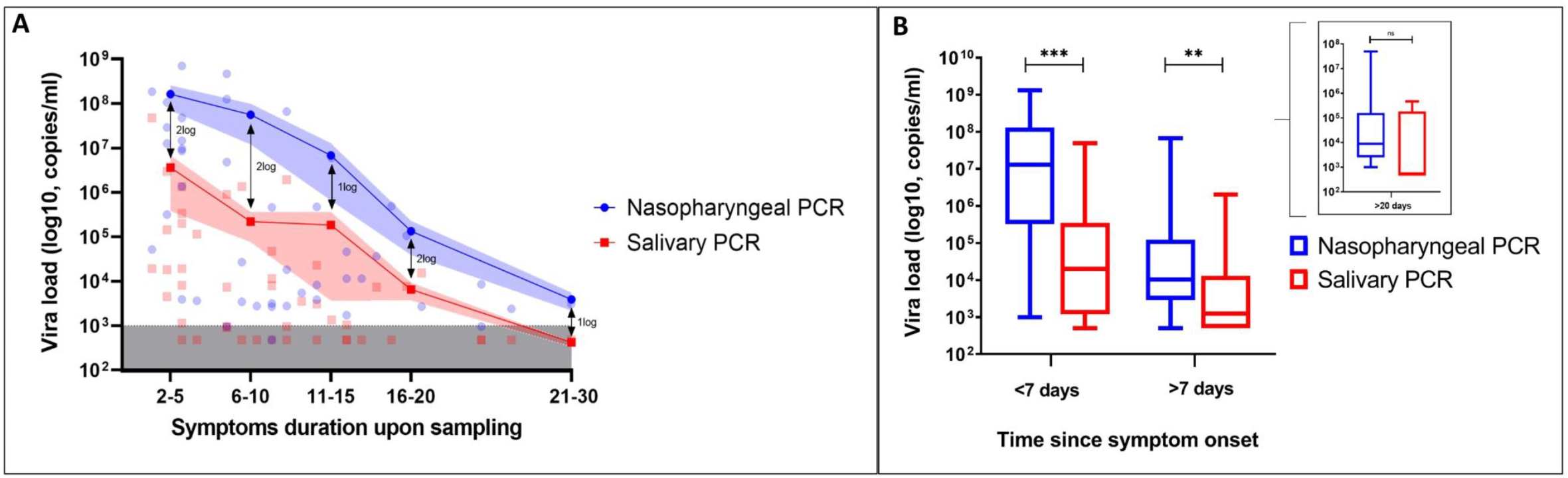
Viral load dynamics of NP RT-PCR versus saliva RT-PCR. **A**. Kinetics of nasopharyngeal and salivary viral load according to symptoms duration upon sampling. The lines connect the mean values of each period and the shaded areas indicate the mean standard errors (SEM). The inferior grey shaded area represents the detection limit (1000 copies/ml). Specimens with undetectable viral load are shown within the grey dashed area. **B**. Box and whisker plots comparing viral load between nasopharyngeal and salivary PCR on different periods since symptom onset. Boxes extend form 25^th^ to 75^th^ percentiles and whiskers show 5 and 95 percentiles. The lines in the middle of the boxes are plotted at median values. (Results were compared using Wilcoxon matched-pairs non parametric test, ***= p<0.001, ** =p<0.01, ns= non significant).

**Figure 3:**
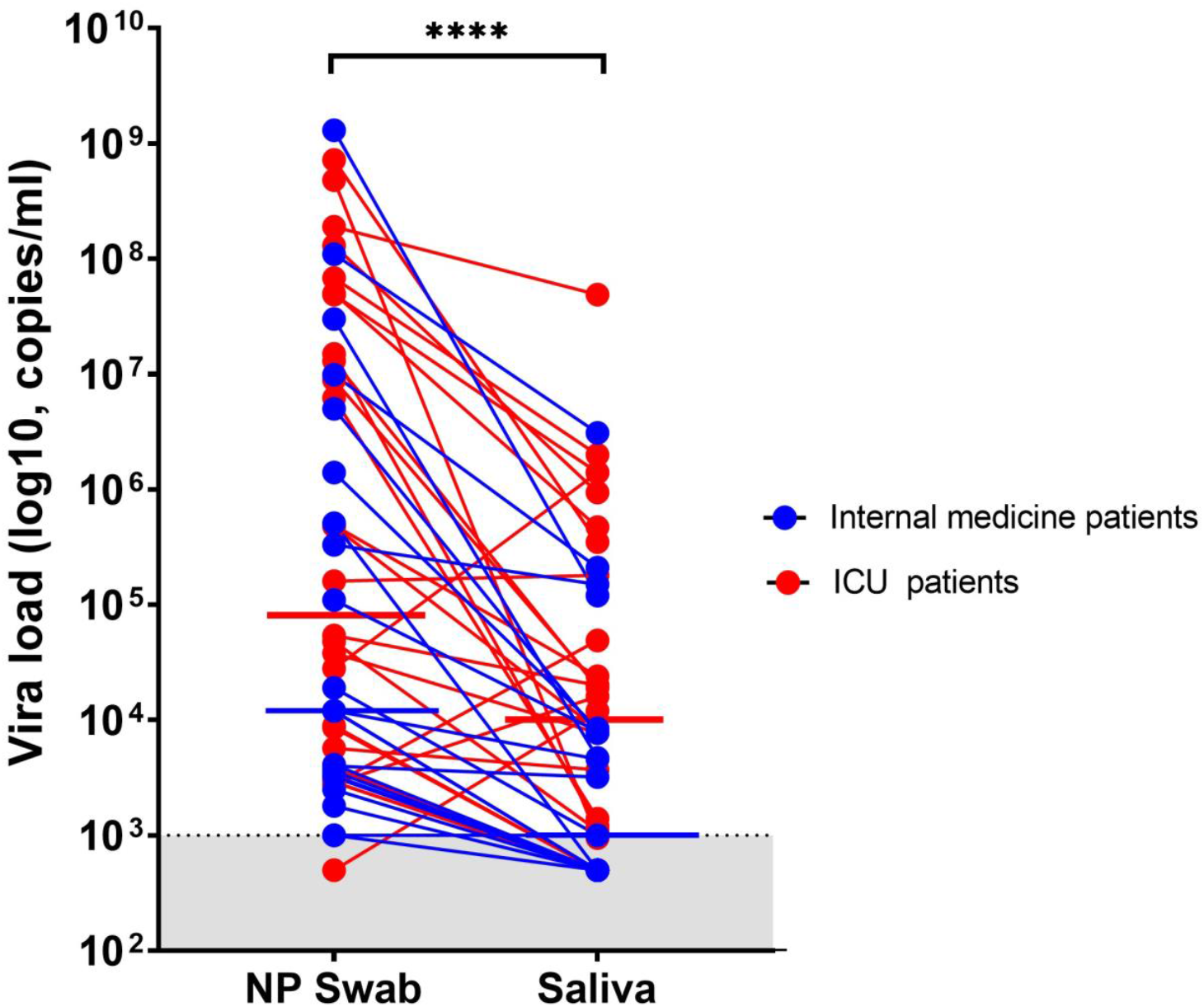
Viral load trend in person-matched NP and saliva samples (n=49). The dotted line and shaded grey area delimit our assay’s detection limit. For graphical representation purposes, samples within the undetectable area are represented with values determined to be at 500 copies/ml. The red and blue horizontal lines represent median values (****=p<0.0001 with Wilcoxon signed-rank test).

RAT exhibited much lower diagnostic performances with sensitivities of 41% and 35% for the Exdia^®^ and Standard Q^®^ assays, respectively among hospitalized patients when a wet-swab approach was used (**Table 2**). Interestingly, the sensitivity of the dry-swab approach was slightly better [sensitivity 47% (95% CI: 35-62) for Standard Q^®^ assay). These antigen tests exhibited very low sensitivity of 8% and 4% for Exdia^®^ and Standard Q^®^ assays, respectively, when applied to salivary swabs. **Figure 4** shows RAT results according to illness duration and VL. All RAT performed better in high VL or early in the course of the disease (especially if VL >10^6^ copies/ml and illness duration < 10 days) (**supplementary material, S4_Figure 3**).

**Figure 4:**
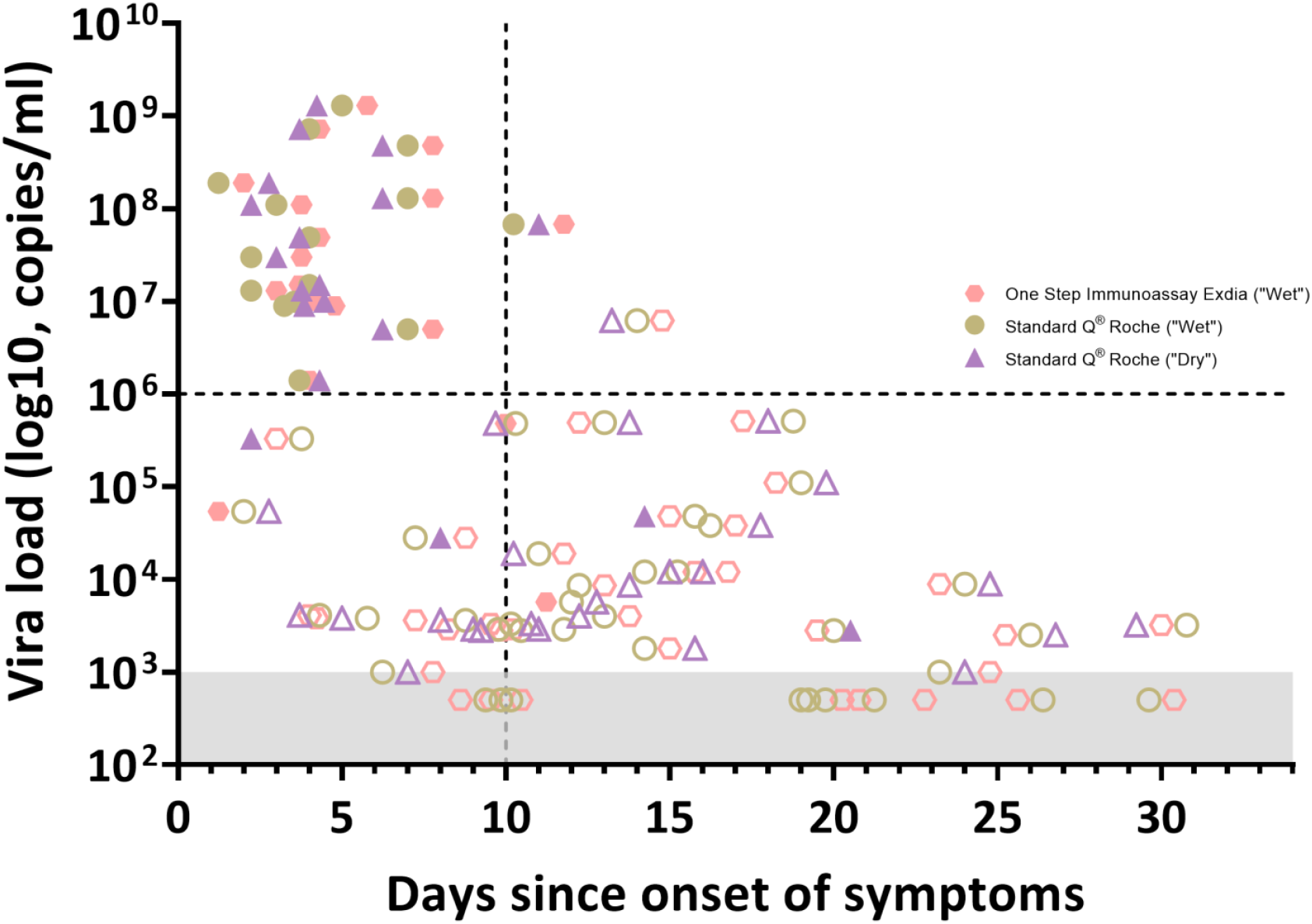
Rapid antigen test results according to time since onset of symptoms and viral load. Full shaded symbols show positive results while no shaded symbols show negative RAT results.

### In vitro dilution effect

The limit of detection for Standard Q^®^ COVID-19 Rapid Antigen Test (Roche-Switzerland) varied between 25 and 27.4 Ct in the dry and wet procedures, respectively, while for Exdia^®^ it varied between 22.2 and 24.7 Ct, respectively, considering the gene E detected by Cobas 6800^®^ (**supplementary material, S5_Table 1**). Quantification of the VL showed lower Ct values (between 2 and 5 Ct corresponding to a difference of 1-2 logs) for the dry series compared to the respective wet series. Ct results from the E or the RdRP gene were comparable, demonstrating a limited intra-method variability.

## Discussion

The present study sought to evaluate the role of alternative and non-invasive methods for diagnosis of SARS-CoV-2 infection in moderately and critically ill-hospitalized patients. Moreover, to our knowledge this is the first study to evaluate the impact of VTM on RAT results for SARS-CoV-2 diagnosis.

Saliva sampling is a promising alternative to NP swabs for SARS-CoV-2 diagnosis, given the ease of collection and comfort for repetitive testing as well as highly reliable results (2-6, 9-12). Nonetheless, only few studies evaluated the diagnostic performance of saliva as compared to NP swab in hospitalized patients (3, 11-13), with results being conflicting. Our study evaluated hospitalized patients with moderate to very severe disease. Most of them were still symptomatic upon sampling (84%) and had a wide distribution of VL, ranging from 10^3^ to 10^9^ copies/ml. Overall, NP RT-PCR was more sensitive in diagnosis of SARS-CoV-2 infection than RT-PCR performed in saliva sample. VL detected in NP swab was higher (1-2 log copies/ml) than in saliva specimens. It is noteworthy however, that sensitivity of salivary RT-PCR increased considerably (from 69% to 81%) when considering patients presenting early in the course of the disease (<10 days). Progressive decrease in VL over time (14) might explain loss of sensitivity of the salivary RT-PCR, which is more pronounced when testing patients with more than 10 days of symptoms. In fact, patients in our study with a positive NP swab and negative saliva sample had low VL (median value of 3700 copies/ml) and a median duration of symptoms of 15 days. This raises the question of the infectivity of those patients and suggest that patients not detected by salivary RT-PCR are those who have low infectivity potential and are late presenters in the course of the disease. Therefore, the results of our study, along with previous published data (3, 9, 11-13), suggest that saliva specimens could be a fair non-invasive alternative to NP swabs (if NP swab is contraindicated for example), especially for those presenting early in the course of illness.

A second goal of our study was to evaluate the diagnostic performance of RAT among hospitalized patients. RAT exhibited unacceptable overall diagnostic performances for hospitalized patients whether performed in NP swab or in saliva specimens. To date, both the Federal Office of Public health in Switzerland and the Swiss Society of Microbiology recommend RAT only within the first 4 days of symptoms (22). Nevertheless, very few (if at all) of hospitalized patients present within this timeframe since the onset of symptoms. When we tested the diagnostic performance of RAT for patients presenting with less than 10 days of illness, sensitivity remained very low. RAT yielded high diagnostic performances only for patients with high VL (≥10^6^ copies/ml) (**supplementary material, S4_Figure 3**). The much lower diagnostic performances of RAT in saliva might be explained by the lower VL in saliva as compared to NP swabs or eventually the presence of mucosal secretory immunoglobulins targeting SARS-CoV-2 antigens and thus competing with RAT for the same target (14, 23). Finally, our study evaluated the role of VTM in SARS-CoV-2 diagnosis by RAT. While both “Wet” and “Dry” procedures are recommended by the manufacturer, previous data suggest that VTM can influence diagnostic performances of RT-PCR and RAT for SARS-CoV-2 detection (15, 24). This is the first study to our knowledge that evaluates “head to head” a “Wet” versus “Dry” RAT procedure for SARS-CoV-2 detection. Our in vitro evaluation showed higher Ct levels for the “Wet” series of both antigen tests used, suggesting a dilution effect when the swab is immerged in the VTM. Our “head to head” clinical comparison confirmed the *in vitro* experimentation, showing that the “Dry” NP swab performed slightly better than the “Wet” one (sensitivity increasing from 35% to 47%), likely due to a decreased dilution of the sample. While RAT can not be recommended for hospitalized patients, the observed difference between “Wet” and “Dry” swabs should be taken into account when performing RAT for SARS-CoV-2 diagnosis in an outpatient setting. Still, when RAT are just used as a supplementary triaging step at the hospital entry to fasten isolation of highly contagious subjects, the use of a wet swab approach is also acceptable since the lower sensitivity is compensated by the fact that in high pandemic period, performing a single sampling for both RAT and PCR is likely a good and effective option (14).

Our study has the strength to test prospectively hospitalized patients with variable disease severity (ICU versus Internal Medicine ward) and a wide distribution of VL and symptoms duration. This study gives an insight on the diagnostic performances of different tests in real life conditions in hospitalized patients. Moreover, it is the first study to our knowledge, to evaluate the impact of VTM on the RAT ability to detect SARS-CoV-2.

On the other hand, this study has a few limitations as well. Its monocentric nature and limited sample size require our results to be confirmed by larger prospective trials. Our patients were initially diagnosed with NP swab RT-PCR that might have induced a bias towards subsequent NP swabs being more often positive versus other samples. Hospitalized patients may have altered saliva production or composition (25) that could influence saliva based diagnostic strategies or even explain the differences observed in salivary RT-PCR performances among severely and mildly ill COVID-19 patients. We chose to use a validated and easy to use non-invasive saliva collection procedure (5). It is possible that other methods of saliva collection (such as throat washing for example) would have improved diagnostic yield and should therefore be tested in other comparative trials.

In conclusion, NP swab RT-PCR was the most sensitive method to diagnose SARS-CoV-2 infection in moderately to critically ill hospitalized patients. Salivary RT-PCR could be used as an alternative non-invasive method if NP swabs are contraindicated, particularly for patients presenting early in the course of the disease. VTM induced dilution effect can impact diagnostic performance of RAT.

## Data Availability

Data will be available upon reasonable request to the authors.

## Contributions

AK, GC and GG conceived and designed the study, performed the literature research, analyzed data and performed statistical analyses, drafted and revised the article. AK and GC collected moreover patient’s data and performed hands-on conduct study’s diagnostic procedures. RB conceived and designed the study, provided essential reagents and materials and performed in vitro experiments. JPM was responsible for project administration in ICU and collected patient’s data. SAM and SG were responsible for project administration in ICU. OO designed the study and provided essential reagents and materials. AC conceived and designed the study and provided essential reagents and materials. PV designed the study and was responsible for the project administration in Internal Medicine Ward. PAB designed the study and was responsible for the project administration in Internal Medicine Ward. JDC designed the study and was responsible for the project administration in ICU. All authors revised the article and approved the submitted version.

## Ethical Approval

The project received approval from the Ethics Committee of canton Vaud, Switzerland (2020– 02818).

## Acknowledgement

We would like to thank all the technical staff of the Microbiology Institute of Lausanne University Hospital who performed routine RT-PCRs as well as the entire medical and nurse team of intensive care unit and internal medicine ward for their precious help in realization of the project.

## Abbreviations

NP: Nasopharyngeal
RT-PCR: Real time polymerase chain reaction
POCT: Rapid Point-of-Care Test
RAT: Rapid Antigen Test
VTM: Viral Transport Medium
ICU: Intensive Care Unit
FOPH: Federal Office of Public Health
VL: Viral Load
Ct: Cycle threshold
IQR: Interquartile range

