## Supplemental File 1 for "Sensitivity of rapid antigen testing and RT-PCR performed on nasopharyngeal swabs versus saliva samples in COVID-19 hospitalized patients: results of a prospective comparative trial (RESTART)"

CHUV
centre hospitalier universitaire vaudois
Institut de Microbiologie

### Procedure for collecting saliva for COVID-19 saliva test (RESTART Study)

#### Before the test:

Make sure the patient will not eat, drink, brush his/her teeth or smoke at least 20 minutes (ideally 1 hour) before taking the sample.

#### How to perform the test:

1. When you are ready to perform the test take the cotton swab out of its pouch.
2. Rub with the end of the cotton swab and for at least 20 seconds the inside of patient's mouth by following the following sequence: internal surface of the cheeks on both sides → upper and then lower gingiva→over- and under- the tongue→ hard palate. Make sure you have put enough saliva on the cotton swab.

#### After the test:

1. Once the sample has been taken, place the swab in the tube, locating the breakable area (**figure 1A**).
2. Break the swab at the breakable part (**figure 1B**).
3. Close the cap properly and shake the tube 2-3 times to mix the sample well with the liquid in the tube (**figure 1C**).
4. Place the protected tube in the plastic sleeve (**figure 1D**), then close it and send it to Microbiology laboratory with patient's details and the service's label (**figure 1E**).

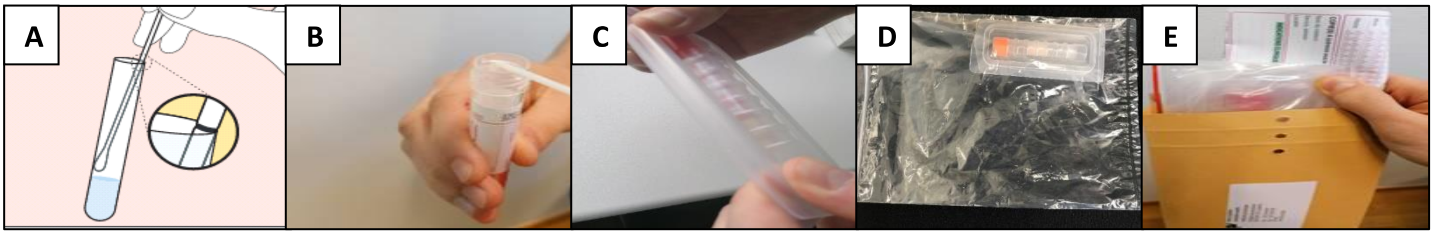


**Figure 1:** Sending sample to microbiology laboratory procedure.

If you have any questions, please contact the COVID microbiology line on 021/314 59 99 (8 am-10pm, 7 days a week)
