## Supplementary figures and images for "Sensitivity of rapid antigen testing and RT-PCR performed on nasopharyngeal swabs versus saliva samples in COVID-19 hospitalized patients: results of a prospective comparative trial (RESTART)"

### Supplemental Figure 1

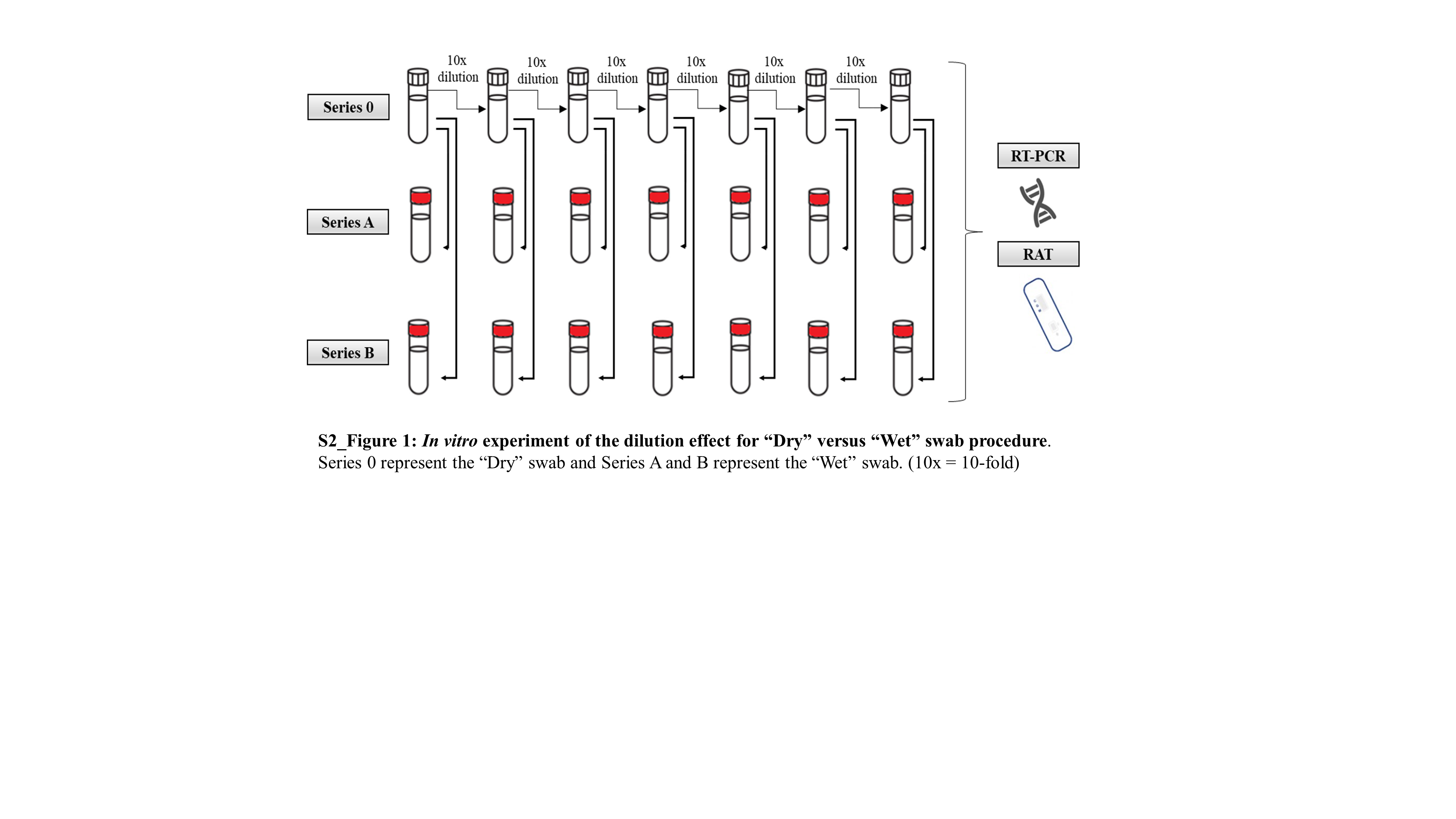

### Supplemental Figure 2

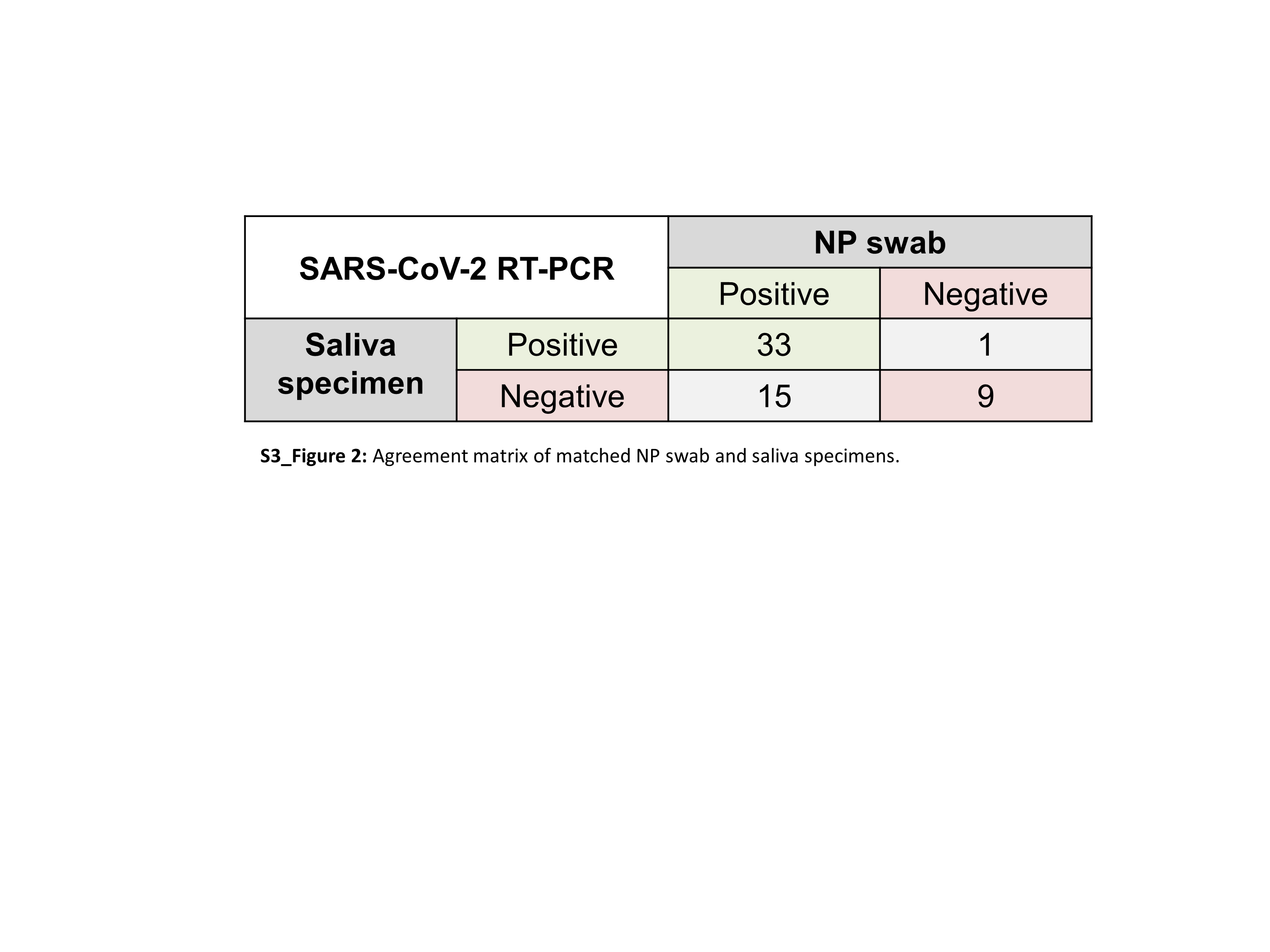

### Supplemental Figure 3

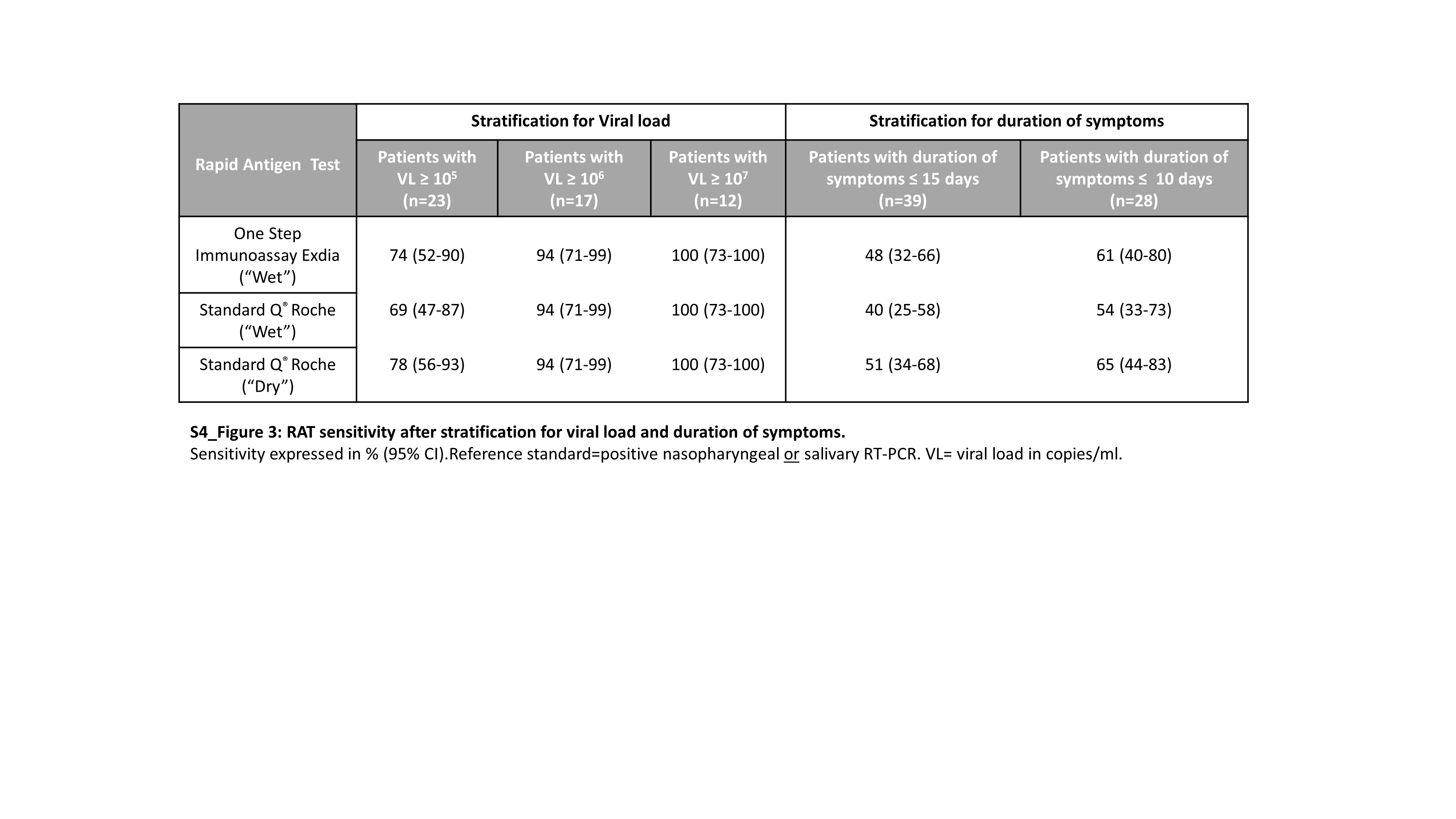
