## Supplemental Table 1 for "Sensitivity of rapid antigen testing and RT-PCR performed on nasopharyngeal swabs versus saliva samples in COVID-19 hospitalized patients: results of a prospective comparative trial (RESTART)"

| **Type of swab** | **Type of test** | **Dil.1** | **Dil.2** | **Dil.3** | **Dil.4** | **Dil.5** | **Dil.6** | **Dil.7** |
| --- | --- | --- | --- | --- | --- | --- | --- | --- |
| “Dry” – Series 0 | One Step Immunoassay Exdia® | P | P* | N | N | N | N | N |
|  | Standard Q^®^ Roche | P | P | P | N | N | N | N |
|  | Cobas 6800  (Ct gene RDRP) | 19.5 | 22.3 | 24.8 | 27.9 | 30.8 | 33.2 | 35.0 |
|  | Cobas 6800  (Ct gene E) | 19.3 | 22.2 | 25.0 | 27.7 | 30.8 | 33.3 | 35.7 |
| “Wet” –  Series A | One Step Immunoassay Exdia® | P* | N | N | N | N | N | N |
|  | Standard Q^®^ Roche | P | P | N | N | N | N | N |
|  | Cobas 6800  (Ct gene RDRP) | 24.9 | 27.6 | 30.7 | 32.9 | 35.4 | 36.7 | 37.8 |
|  | Cobas 6800  (Ct gene E) | 24.7 | 27.4 | 30.8 | 33.2 | 35.5 | 39.5 | 39.6 |
| “Wet” –  Series B | One Step Immunoassay Exdia® | P* | N | N | N | N | N | N |
|  | Standard Q^®^ Roche | P | N | N | N | N | N | N |
|  | Cobas 6800  (Ct gene RDRP) | 24.4 | 27.5 | 30.4 | 33.0 | 34.7 | 37.1 | N |
|  | Cobas 6800  (Ct gene E) | 24.3 | 27.3 | 30.4 | 33.3 | 35.0 | 39.7 | N |

**S5_Table 1:** **Limit of detection (LoD) for dry” versus “wet” swab procedure during the *in vitro* simulation experiment.**

Series 0 was considered the internal reference for LoD, as if it corresponded to a dry swab. Series A and B were obtained after inoculating a swab, previously immerged in the corresponding tube from series 0, inside VTM tubes, thus simulating the process of wet swab. RT-PCR with Cobas 6800 was used as molecular confirmation and Ct calculation (Dil.= dilution; P= Positive test; N= Negative test; *= low positivity).
